# ASSOCIATION BETWEEN VITAMIN D LEVEL AND SERUM LIPID PARAMETERS AMONG INDIAN ADULTS WORKING IN IT SECTOR: A RETROSPECTIVE OBSERVATIONAL STUDY

**DOI:** 10.1101/2020.10.20.20215624

**Authors:** Bhavani Anantharamakrishnan, Jisha Benansia

## Abstract

**Background & objectives:** Association between serum vitamin D levels and lipid levels has been focus of research in recent times. The relationship of 25(OH)D and Atherogenic Index of Plasma is not well established among Indian population. We investigated the prevalence of vitamin D deficiency, its relation with serum lipids and AIP among IT employees.

**Methods:** A retrospective observational study was conducted among employees of multiple IT firms in Bangalore, India. Participants were selected by convenient sampling from annual comprehensive health screening at work place. Serum lipid levels were assessed auto-analyser (ADVIA1650; Siemens, NY, USA). Serum 25(OH)D was assessed by Chemiluminescent microparticle immunoassay. AIP was calculated as log [Triglycerides/HDL-Cholesterol].

**Results:** Among 533 subjects included final analysis, mean of age was 33.18±6.6 with Male: female ratio of 1.8:1. Vitamin D deficiency and AIP > 0.15 was observed in 405 (75.98%) and 450 (84.43%) participants respectively. Median HDL-cholesterol was significantly lower (42 Vs 45.5, P value 0.004) and LDL cholesterol was significantly higher (90 Vs 52, P value <0.001) among people with vitamin-D deficiency. LDL (Spearman Rho=-0.18, P <0.001) had weak negative correlation, Total cholesterol (Spearman Rho=-0.1, P Value <0.01) and VLDL (Spearman Rho=-0.18, P Value <0.001) had weak positive correlation with 25(OH)D. No statistically significant association was observed between AIP and vitamin-D.

**Conclusions:** More than two thirds of IT employees were deficient in vitamin D. Vitamin D deficiency was associated with significantly higher LDL, lower HDL and VLDL levels. No statistically significant association was observed between vitamin D and AIP.

## INTRODUCTION

Vitamin D deficiency had emerged as a global public health concern and is highly prevalent even in tropical nations.[1] In India, studies conducted on different population groups have reported 70% to 90% of the people to be vitamin D deficient. [2-5] Recent evidence showed that low vitamin D level is associated with higher risk of numerous chronic diseases, such as atherosclerosis[6], hypertension[7], cardiovascular diseases [8], diabetes [9],obesity[10], myocardial infarction[11], and stroke[12].

Association between vitamin D deficiency and serum lipids is another area which had attracted attention of researchers across the globe. This assumes huge clinical importance, as dyslipidaemia is an independent risk factor for multiple cardiovascular and cerebrovascular complications [13]. Many researchers documented that individuals with adequate serum vitamin D levels to have more favourable lipid profile compared to people with vitamin D deficiency. A systematic review conducted among children and adolescents reported higher 25(OH)D is to result in favourable lipid profile. This indicates this interrelationship of vitamin D and lipids to be operational from the early ages of an individual’s life[14]. A longitudinal study of adults close to one and half decade revealed an inverse association between serum triglyceride and serum 25(OH)D levels[15]. Vitamin D deficient people also reported to have higher levels of low-density lipoprotein cholesterol by numerous researchers, but the findings were often inconsistent[13] The findings on direction and magnitude of association with other lipid parameters are also conflicting[16]. Few studies also reported gender differences in the pattern of association [16-18].

Atherogenic Index of Plasma (AIP) has been proposed as more accurate biomarker associated with multiple chronic diseases and cardiovascular complications [19, 20]. The association between serum vitamin D level and AIP has been explored by very few studies. A significant negative association among men, with no association among women was reported by a recent study[17]. Others studies documented relatively stronger association in men [21]or significant negative association in both genders [22].

Considering the possible protective role of serum 25(OH)D in maintaining the favourable lipid profile and possible adverse impact of vitamin D deficiency on lipid profile and AIP it is vital to understand this association. Considering the role of various demographic and lifestyle related parameters mediating this association, studies on local population are essential. Hence, we assessed serum 25(OH)D levels, prevalence of vitamin D deficiency and association of serum lipid levels, AIP with vitamin D levels among adults working IT sector.

### Methods

The study was a retrospective observational study conducted among adult population undergoing regular annual health screening from multiple IT firms located in Metropolitan city of Bangalore, India. The final analysis included 533 subjects. Assuming the HDL cholesterol level as the primary outcome of interest, the observed difference in the primary outcome between the subject with and without vitamin D deficiency had yielded 100% power with 5% two-sided alpha error. The study included the data generated from the annual health screening conducted between June 2018 to July 2019 for adults above 18 years of age, both male and female, whose data on serum vitamin D levels and serum lipid levels was available. Considering the retrospective nature of the study important data regarding the potential confounders like vitamin D/ Calcium supplementation, weight loss treatment, use of lipid lowering drugs etc was not available. Incomplete records, where any data related to vitamin D level or any of the lipid profile parameters missing were excluded from the study.

### Estimation of 25 (OH) D

Chemiluminescent microparticle immunoassay (CMIA) with automated instrument for estimation of 25(OH)D was used. Blood samples were obtained after an overnight fast. Total serum 25(OH)D was measured by a ROCHE Modular analytics170 with commercially available kits. The measurable range was 3.0–70.0 ng/mL, with an inter- and intra- assay variable coefficient of 9–15% and <10%, respectively.

### Estimation of serum lipid profile

Serum lipid profile (including triglycerides, total cholesterol, HDL cholesterol and LDL cholesterol), were measured by an auto-analyser (ADVIA1650; Siemens, NY, USA) with commercially available kits. AIP was calculated based on the formula: log [TG/HDL-C] [17] AIP > 0.15 was considered as abnormal value [17]serum (OH) D value ≤20=ng/mL was considered as vitamin D deficiency [13].

### Ethical consideration

The participants data was securely stored on the authenticated cloud platform hosted on servers located in India. All the institutional mechanisms were in place to ensure security of the data as per the required standards. The data was deidentified, anonymized by the institution and data analysis of the deidentified data was done by the collaborating agency within the premises of the mfine on dedicated and secure platforms belonging to the organizations where required. Waiver of consent was obtained from ethical committee and the study was approved by the Digital Health Research Independent Ethics Committee (approval number: DHRIEC/SAL02/21052020, dated: 21.5.2020)

### Statistical methods

Vitamin D was considered as a primary explanatory variable. Serum lipid profile parameters were considered as primary outcome parameters. Age, gender was considered potential confounders. Quantitative variables were assessed for compliance with normal distribution, by visual inspection of histograms, normality Q-Q plot and Shapiro Wilk test p-values. Normally distributed numeric variables were compared between vitamin D deficient and normal subjects using unpaired t-test, Mann Whitney U test was used to compare the Non-normally distributed numeric variables. Chi square/Fisher exact test was used to compare categorical variables. Correlation between serum 25 (OH)D and lipid parameters was assessed by Spearman rank correlation coefficient. P value < 0.05 is considered statistically significant. IBM SPSS version 22 was used for statistical analysis [23].

## RESULTS

A total of 533 subjects were included in the final analysis. The mean of age was 33.18 ± 6.6. The male to female ratio was 1.8:1. Mean serum 25 (OH)D level was 15.69 ± 11.98. Vitamin D deficiency was observed in 405 (75.98%) participants and 450 (84.43%) had AIP > 0.15 The mean values serum lipid parameters are presented in Table 1.

**Table 1:**
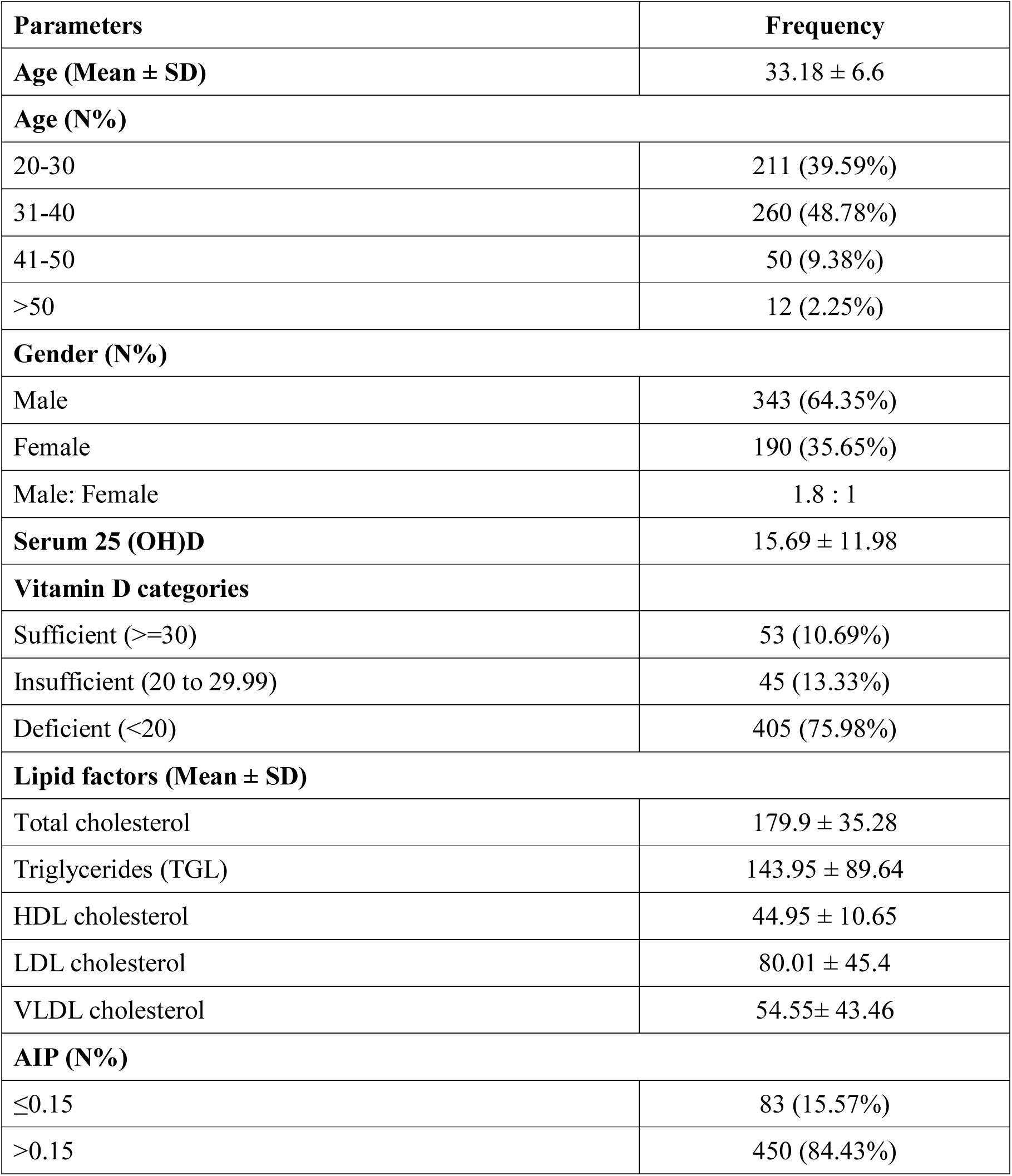
Descriptive analysis of demographic variables in the study population (N=533)

Median HDL cholesterol was significantly lower (42 Vs 45.5, P value 0.004) and LDL cholesterol was significantly higher (90 Vs 52, P value <0.001) among people with vitamin D deficiency. Total cholesterol and VLDL were significantly lower among people with vitamin D deficiency. No statistically significant difference was observed ins total cholesterol level between people with and without vitamin D deficiency. The proportion of people with AIP >0.15 was almost similar in both the groups (84.69% Vs 83.59%, P value 0.765). (Table 2) Stratified analysis was done based on gender to assess whether there are any gender wise differences in association between vitamin D deficiency and serum lipid parameters. Similar pattern of higher LDL, and lower HDL and VLDL levels was observed in both the men and women. Though men had comparatively higher proportion of subjects with AIP>0.15, no statistically significant difference in AIP was observed between people with and without vitamin D deficiency in either gender.(table 3)

**Table 2:**
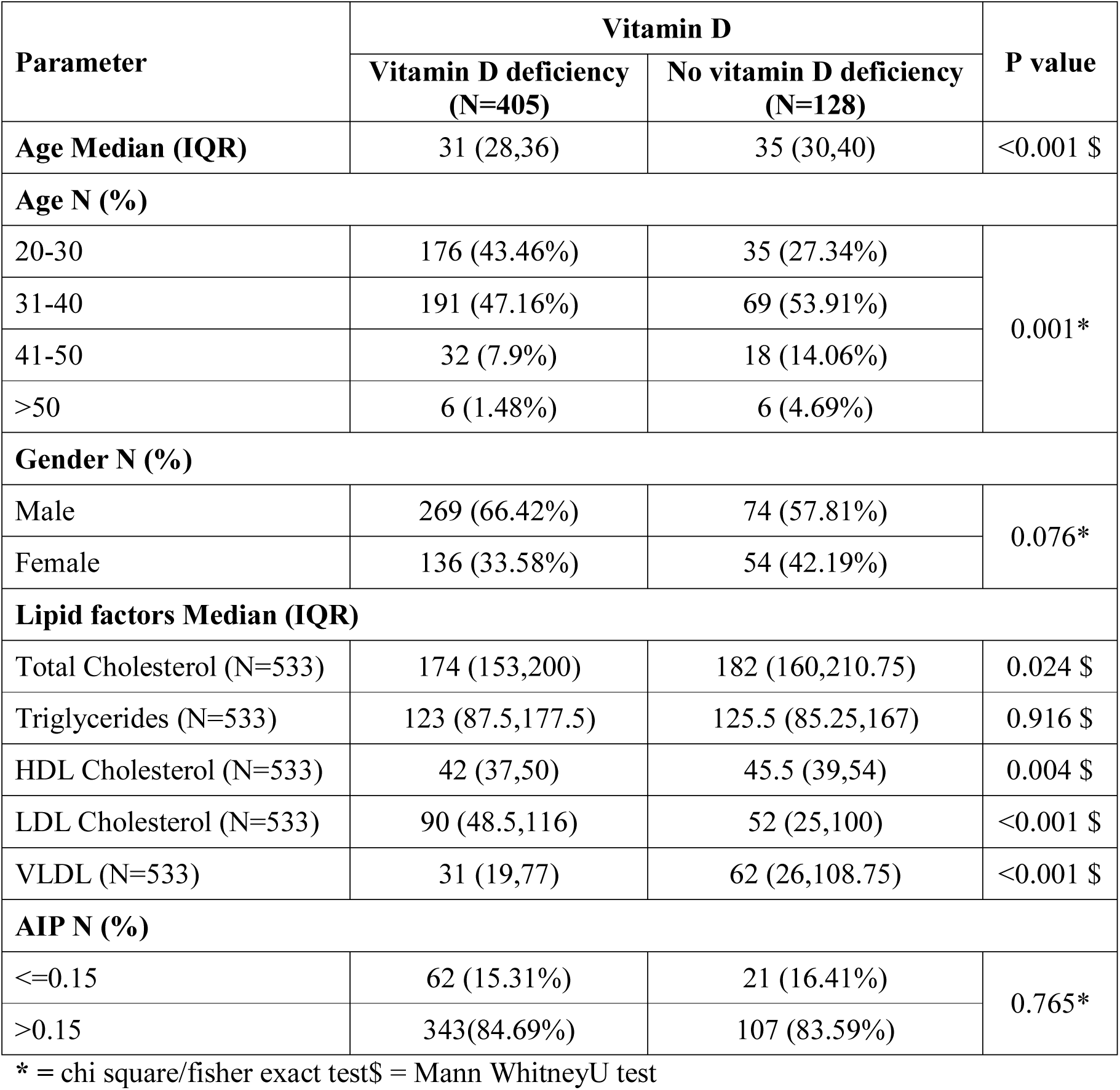
Comparison of demographic and outcome parameters between people with and without Vitamin D deficiency (N=533)

**Table 3:** Gender stratified comparison of serum lipid parameters between Vitamin D deficiency groups (N=533)

| Parameter | Male |  |  | Female |  |  |
| --- | --- | --- | --- | --- | --- | --- |
|  | Vitamin D deficiency | No vitamin D deficiency | P value | Vitamin D deficiency | No vitamin D deficiency | P Value |
| **Lipid factors, Median (IQR)** |  |  |  |  |  |  |
| Total Cholesterol | 181 (159.5,208) | 189 (173.25, 214.25) | 0.019 \$ | 164.5 (147, 183.75) | 170 (149, 203.75) | 0.29 \$ |
| Triglycerides | 139 (96,197) | 139.5 (107.25, 187.5) | 0.58 \$ | 96 (69, 135.75) | 99.5 (69.75, 134.25) | 0.98 \$ |
| HDL Cholesterol | 40 (35,46) | 43 (37, 51.25) | 0.025 \$ | 48 (42,55) | 51 (42.75, 61.25) | 0.21 \$ |
| LDL Cholesterol | 93 (42,121) | 55.5 (27, 109.25) | 0.007 \$ | 87.5 (61,105) | 40 (20, 92.25) | 0.001 \$ |
| VLDL | 36 (22,87) | 78.5 (28.75,116) | <0.001 \$ | 24.5 (15.25, 50) | 59.5 (19,103) | 0.001 \$ |
| **AIP, N (%)** |  |  |  |  |  |  |
| <=0.15 | 22 (8.18%) | 6 (8.11%) | 0.984* | 40(29.41%) | 15 (27.78%) | 0.823* |
| >0.15 | 247(91.82%) | 68 (91.89%) |  | 96(70.59%) | 39 (72.22%) |  |

\* = Chi square/fisher exact test, \$ = Mann Whitney test

Among the serum lipid parameters, only LDL had shown statistically significant but weak negative correlation (Spearman Rho=-0.18, P Value <0.001) and VLDL has shown statistically significant but weak positive correlation (Spearman Rho=-0.18, P Value <0.001). Though HDL had shown a strong positive correlation, the correlation was statistically not significant (Spearman Rho=-0.0.70, P Value 0.106). Total cholesterol and Triglycerides have shown no statistically significant correlation with serum 25 (OH) D values. (Figure 1a to 1e)

**Figure 1(a):**
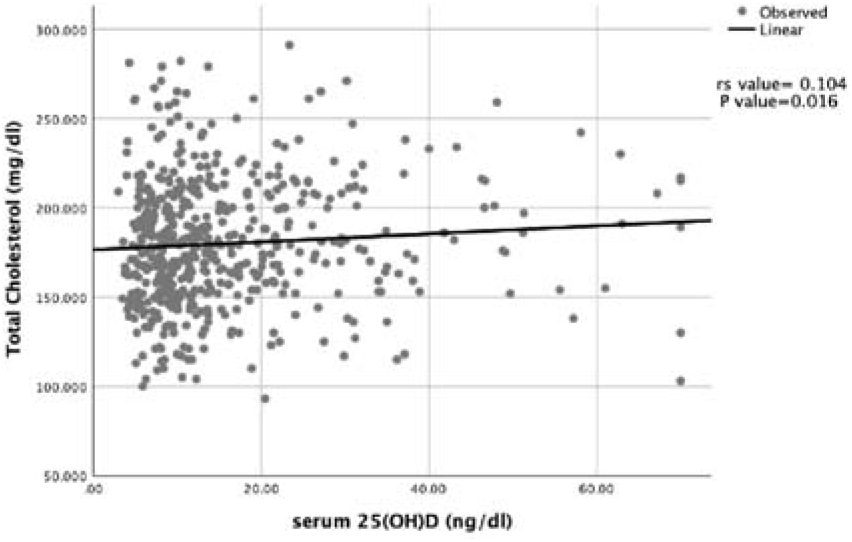
Scatter plot for correlation between total cholesterol and Serum 25(OH)D (ng/dl)in the study population (N=533)

**Figure 1(b):**
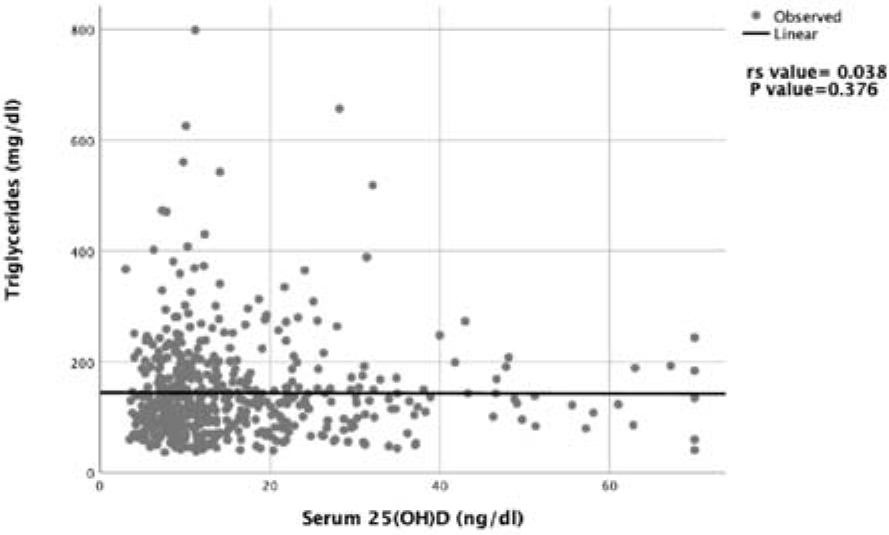
Scatter plot for correlation between triglycerides and Serum 25(OH)D (ng/dl) in the study population (N=533)

**Figure 1(c):**
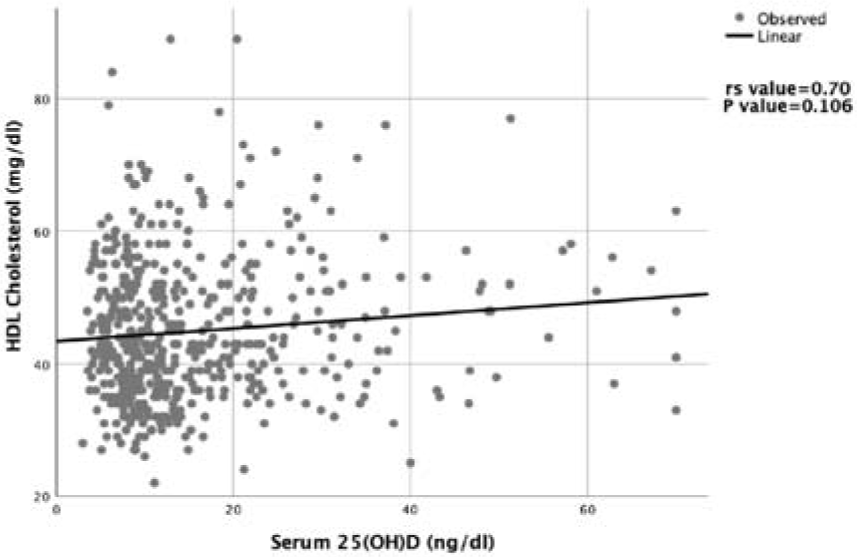
Scatter plot for correlation between HDL and Serum 25(OH)D (ng/dl) in the study population (N=533)

**Figure 1(d):**
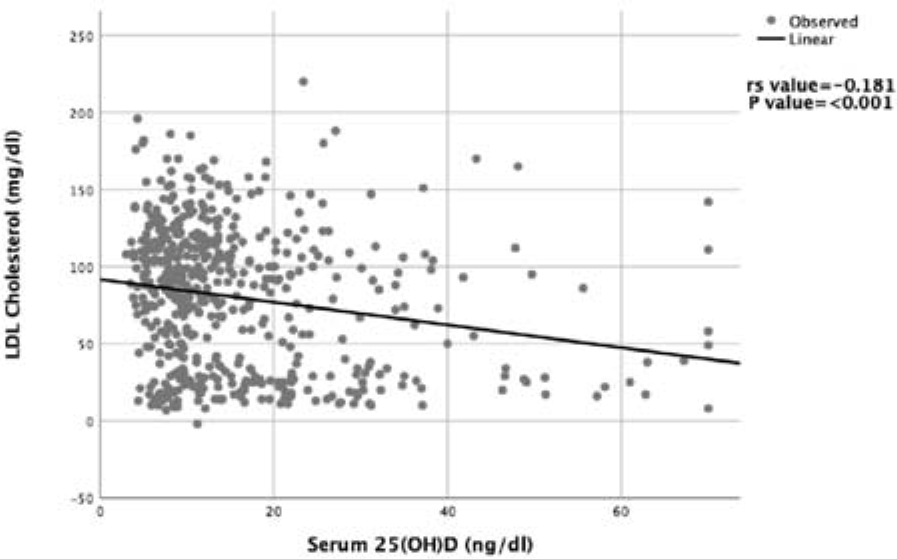
Scatter plot for correlation between LDL and Serum 25(OH)D (ng/dl) in the study population (N=533)

**Figure 1(e):**
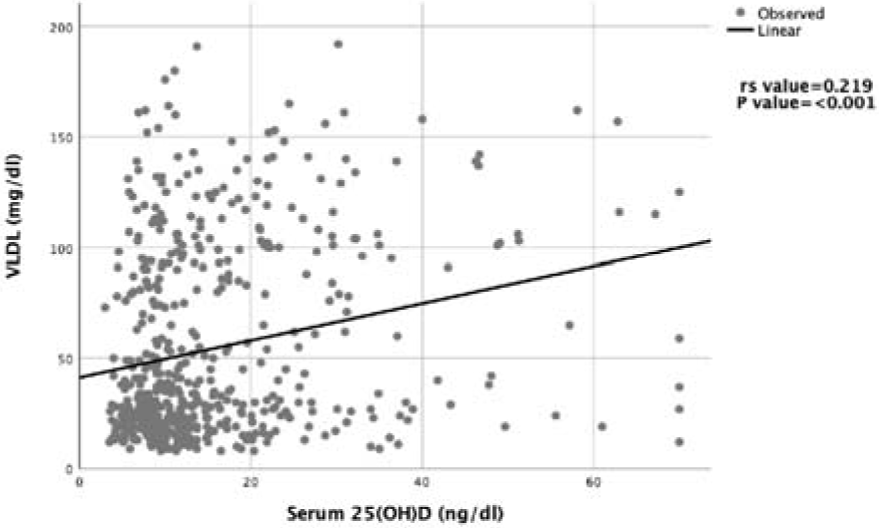
Scatter plot for correlation between VLDL and Serum 25(OH)D (ng/dl) in the study population (N=533)

**Figure 1(f):**
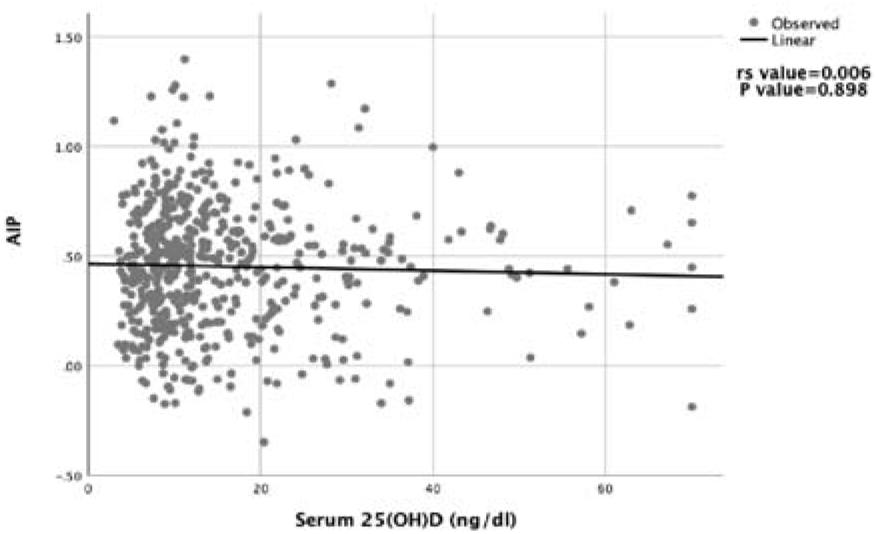
Scatter plot for correlation between AIP and Serum 25(OH)D (ng/dl) in the study population (N=533)

## DISCUSSION

Insufficient vitamin D levels may contribute to many chronic diseases in addition to adverse bone health. Low vitamin D is found to have an unfavourable effect on body lipids, since it may increase intestinal absorption and synthesis of lipids, reduce lipolysis and lipid metabolism, and reduce secretion and sensitivity of insulin and thereby increase the risks of diabetes, metabolic syndrome and cardiovascular disease [24]. However, the evidence is limited.

In the present study, the overall vitamin D deficiency was seen in 75.98% population. Reported prevalence of vitamin D deficiency was quite varied across the studies. Wang, Y., et al [17]. in their study found that the overall vitamin D deficiency seen was 58.5% (males 54.4%, females 63.7%). In an Indian study by Chaudhuri, J. R., et al [13]Vitamin D deficiency was observed among 39.3% of study subjects. In a cross-sectional study Kazak adults Zhang, M. C., et al[25]reported 72.4% of participants to be vitamin D deficient. Among a group of premenopausal women Patel, P. A., et al[26]have reported a 93% prevalence of vitamin D deficiency, indicating possible role in osteoporosis and increased risk of cardiovascular morbidity post menopause. Sun, X., et al [27] showed that the deficiency was 78.7% in Japanese men. These differences can be attributed to differences study population, ethnicity, gender, sun exposure, occupation and dietary intake and highlights the need for the studies on specific local population groups to assess the true burden.

In the present study people with vitamin D deficiency had significantly elevated LDL cholesterol levels, whereas TC, VLDL and HDL were found to significantly lower among vitamin D deficient individuals. Similar pattern was observed both in males and females. In our study, serum 25 (OH)D levels have shown statistically significant but weak negative correlation with LDL. The pattern of association reported between 25 (OH)D level and lipid parameters is quite heterogenous across the studies. Similar to our study Lupton, J. R., et al[28] reported people with deficient serum 25(OH)D to have significantly lower serum HDL-C (−5.1%) and higher directly measured LDL-C (+13.5%). But the direction of association observed with other parameters like TC and VLDL was not in line with current study findings. Serum 25(OH)D levels were inversely associated with TG and LDL-C, but positively associated with TC among Chinese men[17]. Jungert, A., et al[18] found that 25(OH)D3 was associated with HDL-C (beta=0.197), LDL-C:HDL-C (beta=-0.298) and TC:HDL-C (beta=-0.302) in women and not in men. As per Sun, X., et al [27] serum 25(OH)D concentration was inversely related to the LDL-C/HDL-C and Triglycerides, after controlling for all potential confounding variables. Serum vitamin D had a statistically significant inverse association with TC, LDLC, TG, and homocysteine but had shown positive association with HDLC among hyperlipidaemic individuals [29]. Karhapaa, P., et al[30]found a significant inverse association of 25(OH)D with TC, LDL-C and TG (Beta coefficient of 0.15, 0.13 and 0.17respectively). In contrast to current study, no association was found with HDL-C.

AIP has been proposed as a superior marker of atherogenicity and cardiovascular complications by many researchers, then individual lipid profile parameters [19]. We have observed no statistically significant correlation between serum 25 (OH) D levels and AIP. Wang, Y., et al [17]have reported a weak but statistically significant negative correlation with between serum 25(OH)D concentrations and AIP in men (r = −0.111, p < 0.01) but not in women. Inc contrast Izadi, A., et al[22] found a strong negative correlation between vitamin D levels and AIP. The differences in the demographic composition of the study participants and the prevalence of other co-morbidities would have contributed to the reported heterogenicity across the studies.

The key limitation in our study was the use of a retrospective records and convenience sample affecting the external validity. Also, our study population included only the IT population which is literate and belongs to a higher socioeconomic stratum, which also reduces the ability to generalize the findings. Another potential limitation of our study was our inability to determine the physical activity levels, sunlight exposure, dietary habits, nutritional intake and general lifestyle habits of the included population. Inability to adjust for confounding due these variables is another key limitation. Despite the above limitations, our study established that large proportion of Indian population working in IT sector could be vitamin D deficient and vitamin D may have influence on serum lipid levels.

## CONCLUSIONS

The study established that more than two thirds of adults working IT sector are deficient in vitamin D. People with Vitamin D deficiency tend to have higher LDL and lower HDL cholesterol levels. No statistically significant association was observed between vitamin D and AIP. All the statistically significant correlations observed were weak. Considering the conflicting reports, large multicentric studies among different population subgroups may aid us in better understanding the relationship between vitamin D and serum lipid profile.

## Data Availability

The study included the data generated from the annual health screening conducted between June 2018 to July 2019 for adults above 18 years of age, both male and female, whose data on serum Vitamin D levels and serum lipid levels was available.

## Acknowledgements

We acknowledge the professional support by Evidencian Research associates in data analysis and manuscript editing.

## Declaration of Conflict of Interest

The authors declare there are no conflict of interests.

## Funding Statement

The project was not supported by any extramural fund.

## Notes

### Competing Interest Statement

The authors have declared no competing interest.

### Clinical Trial

A retrospective observational study

### Funding Statement

No external funding was received

### Author Declarations

Digital Health Research IEC(DHRIEC)

